# Impact of Race, Gender, and Insurance Status on Outcomes Following Endovascular Treatment for Acute Ischemic Stroke

**DOI:** 10.1101/2025.04.09.25325561

**Authors:** Tarlan Kehtari, Mona Roshan, Grayson Gigliotti, Chrisnel Lamy, Starlie Belnap, Italo Linfante, Guilherme Dabus

## Abstract

**Background:** Racial, gender, and socioeconomic disparities in stroke outcomes are well documented, but their impact on procedural success and clinical outcomes after endovascular treatment (EVT) for acute ischemic stroke remains unclear.

**Methods:** We retrospectively analyzed 584 acute ischemic stroke patients who underwent EVT (2016-2023), evaluating procedural reperfusion (TICI 2B-3), stroke severity (NIHSS/mRS) at admission and discharge, functional outcomes at discharge and 90 days (mRS), discharge disposition, and thrombolysis (tPA) administration. Multivariable logistic regression assessed independent predictors of outcomes.

**Results:** Successful reperfusion was achieved in 90.8%, with no significant differences by race, gender, or insurance status. Black patients and Medicare-insured individuals presented with significantly greater stroke severity (NIHSS ≥9, p<0.05). Poor functional outcomes (mRS 3-6) at discharge and 90 days were significantly higher among Black patients, females, and Medicare-insured patients (p<0.05). Medicare and Medicaid patients were more frequently discharged to non-home settings. Insurance status was significantly associated with lower likelihood of tPA administration (p=0.006). Logistic regression showed that initial stroke severity strongly predicted outcomes, while procedural success was uniform across demographic groups.

**Conclusions:** Procedural success of EVT was equitable; however, disparities persisted in stroke severity at admission and long-term outcomes. These findings highlight the need for systemic interventions addressing pre-hospital care, prevention, and equitable post-stroke rehabilitation access.

## Introduction

Stroke is a leading cause of adult disability and death, and its impact is not felt equally across populations. There are well-documented racial disparities in stroke incidence and outcomes: the risk of a first-ever stroke is nearly twice as high in Black individuals compared to White, and stroke mortality rates are higher in Black populations.^1^ Similarly, gender differences have been observed – women experience a disproportionate stroke burden, partly due to longer life expectancy, and tend to present with more severe neurologic deficits and have worse functional outcomes than men.^1^ Socioeconomic factors, often approximated by insurance status in the U.S. healthcare system, also influence stroke care and outcomes. Uninsured or underinsured patients are less likely to receive preventive care and may have higher stroke severity on presentation; notably, the mortality risk of uninsured stroke patients is reported to be significantly higher than that of insured patients,^2^ and uninsured patients often present with greater neurologic impairment compared to insured patients.^3^ These disparities stem from a complex interplay of factors including differences in risk factor burden, healthcare access, and quality of care across the stroke continuum.^4^

Advances in acute stroke therapy, particularly endovascular treatment (EVT) for large vessel occlusion, have dramatically improved outcomes for many patients.^5,6^ Ensuring that these benefits reach all segments of the population is a public health priority. Prior studies have identified disparities in access to acute stroke treatments: for example, patients from disadvantaged backgrounds are less likely to receive intravenous thrombolysis (tPA) or thrombectomy, even after accounting for eligibility.^7,8^ However, among those who do undergo EVT, the evidence is mixed on whether race, gender, or socioeconomic status affect procedural success or outcomes. Some single-center analyses have found no difference in reperfusion success or clinical outcomes between Black and White patients treated with thrombectomy,^9^ suggesting that outcomes can be equitable when care is received. On the other hand, broader registry studies point to lingering outcome gaps, particularly related to post-stroke care and recovery, in minority and low-income groups.^7,8,10,11^ Few studies have comprehensively examined the combined influence of race, gender, and insurance status on both the technical and clinical outcomes of EVT.

This study aims to address this gap by analyzing a large retrospective cohort of acute ischemic stroke patients who underwent EVT at our institution, focusing on whether patient race, gender, and insurance status are associated with differences in reperfusion rates, neurological improvement, and functional outcomes. We hypothesized that any observed disparities in overall outcomes would be explained more by differences in stroke severity at presentation and post-procedural care rather than differences in the efficacy of the endovascular procedure itself. By integrating newly updated data and analyses, we seek to clarify the roles of these demographic and socioeconomic factors in stroke treatment outcomes and discuss implications for clinical practice and policy to advance health equity in stroke care.

## Methods

### Study Design and Population

We conducted a retrospective analysis of consecutive patients with acute ischemic stroke due to large vessel occlusion who underwent endovascular treatment at a comprehensive stroke center between January 2016 and December 2023. Patients were identified from a prospectively maintained stroke intervention database. Inclusion criteria were: age ≥18 years, clinical diagnosis of acute ischemic stroke, evidence of a large vessel occlusion on vascular imaging, and treatment with endovascular thrombectomy. We excluded patients with incomplete data on key variables (race, insurance, or outcomes) or those who had stroke mimics. This study was approved by the institutional review board with a waiver of informed consent due to its retrospective nature.

### Variables and Definitions

Patient demographic information collected included self-identified race/ethnicity (categorized as White, Black, Hispanic, or Other), gender, and primary insurance status at admission (categorized as private insurance, Medicare, Medicaid, or uninsured). For analysis, we defined “underinsured” as patients with Medicaid or no insurance, given the similar challenges in access and resources for these groups, and contrasted them with those having private or Medicare insurance (considered “insured”). Clinical data included baseline stroke severity measured by the NIHSS on presentation, comorbidities (hypertension, diabetes, atrial fibrillation, etc.), and stroke risk factors. Process of care variables included onset-to-door time, door-to-groin puncture time, procedure duration, and use of intravenous thrombolysis before EVT when applicable.

The primary procedural outcome was successful reperfusion, defined as modified Thrombolysis in Cerebral Infarction (mTICI) grade 2b or 3 at the end of the thrombectomy procedure (indicating reperfusion of ≥50% of the affected territory). Primary clinical outcomes were neurological status at 24 hours and functional outcome at 90 days. Early neurological improvement was evaluated by the change in NIHSS from admission to 24 hours (or at discharge if earlier), and symptomatic intracerebral hemorrhage (sICH) within 36 hours was recorded (sICH defined as parenchymal hematoma on imaging with neurological worsening). Functional outcome was assessed by the modified Rankin Scale (mRS) at 90 days post-stroke, obtained through outpatient follow-up or telephone assessment by certified personnel blinded to the study hypothesis. For functional outcome analyses, we defined favorable outcome as mRS 0–2 (independent functional status) and poor outcome as mRS 3–6. In-hospital mortality and discharge disposition (home, inpatient rehabilitation, skilled nursing facility or hospice) were also recorded as secondary outcomes.

### Statistical Analysis

We stratified the cohort by race (White vs. Black vs. Hispanic vs. Other), by gender, and by insurance category (private, Medicare, Medicaid, uninsured; and additionally insured vs. underinsured as defined above) to compare baseline characteristics and outcomes. Continuous variables were summarized as mean ± standard deviation or median with interquartile range [IQR] as appropriate, and categorical variables as counts and percentages. Group comparisons were performed using Student’s t-test or Mann-Whitney U test for continuous variables and chi-square or Fisher’s exact tests for categorical variables. We used analysis of variance (ANOVA) or Kruskal-Wallis test for multi-group comparisons (e.g., across four insurance categories), followed by post-hoc pairwise comparisons with Bonferroni correction.

To identify independent predictors of outcomes, we constructed multivariable logistic regression models. Three separate models were prespecified: (1) predictors of successful reperfusion (mTICI 2b-3), (2) predictors of favorable 90-day outcome (mRS 0–2), and (3) predictors of in-hospital mortality. Candidate independent variables included demographics (race, gender, insurance group), initial NIHSS, age, time from stroke onset to reperfusion, and clinically relevant comorbidities (e.g., atrial fibrillation for cardioembolic strokes) or treatment variables (IV tPA use). We used backward stepwise selection with a threshold of p<0.10 for inclusion, while forcing the main variables of interest (race, gender, insurance) into the models. Adjusted odds ratios (OR) with 95% confidence intervals were reported. We checked model calibration and discrimination and assessed for multicollinearity among covariates.

All statistical tests were two-sided with a significance level set at p<0.05. Analyses were performed using SPSS version 28 (IBM Corp.) and R version 4.2.

### Data Availability

The data that support the findings of this study are available from the corresponding author upon reasonable request, in accordance with institutional policies and patient privacy regulations.

## Results

### Demographics and Baseline Characteristics

A total of 584 patients were included in the analysis. The gender distribution was 53.25% female (n = 311) and 46.75% male (n = 273). The racial composition was predominantly Latin/Hispanic (64.68%), followed by White (21.0%), Black (11.90%), and Other (2.42%). Regarding insurance status, 60.34% of patients were covered by Medicare, 21.42% by commercial insurance, 9.87% by Medicaid, and 8.38% were uninsured (Table 1).

**Table 1:** Demographic and Baseline Characteristics of the Cohort (n=584)

| Characteristic | Number of Patients (n) | Percentage (%) |
| --- | --- | --- |
| Gender |  |  |
| Male | 273.0 | 46.75 |
| Female | 311.0 | 53.25 |
| Race |  |  |
| Latin/Hispanic | 378.0 | 64.68 |
| White | 123.0 | 21.0 |
| Black | 70.0 | 11.9 |
| Other | 13.0 | 2.42 |
| Insurance Status |  |  |
| Medicare | 352.0 | 60.34 |
| Commercial Insurance | 125.0 | 21.42 |
| Medicaid | 58.0 | 9.87 |
| Uninsured | 49.0 | 8.38 |

### Procedural Reperfusion

Procedural reperfusion (TICI 2B-3) was achieved in 90.77% (n = 423) of patients. There was no significant difference in reperfusion success by gender (p = 0.333), race (p = 0.784), or insurance status (p = 0.8545). Similar findings were observed for a higher threshold of reperfusion (TICI 2C-3), where 62.16% of patients achieved TICI 2C-3 perfusion, with no statistically significant differences by demographic or insurance factors (Table 2).

**Table 2:** Procedural Reperfusion Outcomes (n=584)

| Subgroup | # of Patients Achieving Reperfusion (n) | Percentage (%) | p-value |
| --- | --- | --- | --- |
| **Overall** | 423 | 90.77 | N/A |
| **Male** | 199 | 92.13 | 0.333 |
| **Female** | 222 | 89.52 | 0.333 |
| **Latin/Hispanic** | 263 | 91.0 | 0.784 |
| **White** | 85 | 91.4 | 0.784 |
| **Black** | 45 | 90.0 | 0.784 |
| **Other** | 10 | 100.0 | 0.784 |
| **Medicare** | 246 | 90.44 | 0.8545 |
| **Commercial Insurance** | 84 | 93.33 | 0.8545 |
| **Medicaid** | 39 | 90.7 | 0.8545 |
| **Uninsured** | 34 | 91.89 | 0.8545 |

### Stroke Severity at Admission (NIHSS and mRS at Admission)

Among the cohort, 39.38% (n = 230) of patients had NIHSS scores between 0-8 at admission, while 60.62% (n = 354) had NIHSS scores ≥9. There were significant differences in NIHSS scores by race (p = 0.026), with Black patients presenting with a higher proportion of NIHSS ≥9. Insurance status was also significantly associated with NIHSS at admission (p < 0.001), with Medicare patients having a greater proportion of NIHSS ≥9. Gender differences in NIHSS at admission were not statistically significant (p = 0.097) (Table 3).

**Table 3:** NIH Stroke Scale (NIHSS) Score at Admission (n=584)

| **Subgroup** | **NIHSS 0-8 (Favorable Outcome) (n, %)** | **NIHSS 9+ (Poor Outcome) (n, %)** | **p-value** |
| --- | --- | --- | --- |
| **Overall** | 230 | 354 | N/A |
| **Male** | 107 | 166 | 0.097 |
| **Female** | 123 | 188 | 0.097 |
| **Latin/Hispanic** | 128 | 250 | 0.026 |
| **White** | 45 | 78 | 0.026 |
| **Black** | 27 | 43 | 0.026 |
| **Other** | 5 | 8 | 0.026 |
| **Medicare** | 102 | 250 | <0.001 |
| **Commercial Insurance** | 58 | 67 | <0.001 |
| Medicaid | 34 | 24 | <0.001 |
| Uninsured | 36 | 13 | <0.001 |

Similarly, mRS scores at admission differed significantly by race (p = 0.019) and insurance status (p < 0.001). Overall, 33.90% (n = 198) of patients had an mRS of 0-2 at admission, while 66.10% (n = 386) had an mRS of 3-6. Black patients and Medicare patients had higher proportions of mRS 3-6. Gender differences in mRS at admission were not statistically significant (p = 0.112) (Supplemental Table 1).

### Neurological Outcomes (NIHSS at Discharge)

At discharge, 60.89% (n = 356) of patients had NIHSS scores between 0-8, while 39.11% (n = 228) had NIHSS scores ≥9. NIHSS at discharge was not significantly associated with gender (p = 0.270), race (p = 0.333), or insurance status (p = 0.294) (Supplemental Table 2).

### Functional Outcomes (mRS at Discharge and 90 Days)

At discharge, 22.95% (n = 134) of patients had an mRS of 0-2, while 77.05% (n = 450) had an mRS of 3-6. Gender differences were statistically significant (p = 0.036), with females having a higher proportion of mRS 3-6 compared to males. Race was also significantly associated with mRS at discharge (p = 0.045), with Black patients having a higher proportion of poor functional outcomes. Insurance status was significantly associated with mRS at discharge (p < 0.001), with Medicare patients having higher rates of mRS 3-6 compared to other groups (Table 4).

**Table 4:** Modified Rankin Scale (mRS) Scores at Discharge (n=584)

| Subgroup | mRS 0-2 (Good Outcome) (n, %) | mRS 3-6 (Poor Outcome) (n, %) | p-value |
| --- | --- | --- | --- |
| Overall | 134 | 450 | N/A |
| Male | 73 | 203 | 0.036 |
| Female | 61 | 250 | 0.036 |
| Latin/Hispanic | 80 | 268 | 0.045 |
| White | 31 | 82 | 0.045 |
| Black | 12 | 53 | 0.045 |
| Other | 2 | 11 | 0.045 |
| Medicare | 46 | 277 | <0.001 |
| Commercial Insurance | 52 | 63 | <0.001 |
| Medicaid | 14 | 39 | <0.001 |
| Uninsured | 13 | 32 | <0.001 |

At 90 days, 36.07% (n = 211) of patients had an mRS of 0-2, while 63.93% (n = 373) had an mRS of 3-6. Similar to discharge outcomes, females (p = 0.036), Black patients (p = 0.021), and Medicare patients (p < 0.001) had significantly higher proportions of poor functional outcomes at 90 days (Table 5).

**Table 5:** Modified Rankin Scale (mRS) Scores at 90 Days (n=584)

| Subgroup | mRS 0-2 (Good Outcome) (n, %) | mRS 3-6 (Poor Outcome) (n, %) | p-value |
| --- | --- | --- | --- |
| Overall | 211 | 373 | N/A |
| Male | 108 | 156 | 0.036 |
| Female | 103 | 217 | 0.036 |
| Latin/Hispanic | 134 | 213 | 0.021 |
| White | 48 | 70 | 0.021 |
| Black | 15 | 51 | 0.021 |
| Other | 3 | 10 | 0.021 |
| Medicare | 86 | 234 | <0.001 |
| Commercial Insurance | 62 | 58 | <0.001 |
| Medicaid | 27 | 43 | <0.001 |
| Uninsured | 23 | 38 | <0.001 |

### Discharge Disposition

The majority of patients (83.56%, n = 488) were discharged home, while 16.44% (n = 96) were discharged to hospice or an inpatient facility. Gender differences approached statistical significance (p = 0.077), with males more likely to be discharged home than females. Insurance status was significantly associated with discharge disposition (p = 0.001), with Medicare and Medicaid patients more likely to be discharged to hospice or inpatient facilities. Race was not significantly associated with discharge disposition (p = 0.274) (Supplemental Table 3).

### Intravenous (IV) Thrombolysis Administration

Among the cohort, 40.07% (n = 234) of patients received tPA, while 59.93% (n = 350) did not. Gender (p = 0.931) and race (p = 0.891) were not significantly associated with IV thrombolysis administration. However, insurance status was significantly associated with IV thrombolysis administration (p = 0.006), with Medicare patients less likely to receive IV thrombolysis compared to other insurance groups (Supplemental Table 4).

### Logistic Regression Analysis: Statistically Significant Predictors of Poor Outcomes

Multivariable logistic regression models were used to assess the independent associations of gender, race, and insurance status with procedural reperfusion, stroke severity at admission, neurological outcomes at discharge, functional outcomes, and post-hospital disposition.

Procedural reperfusion success (TICI 2B-3 and TICI 2C-3) was not significantly associated with gender, race, or insurance status, indicating that the likelihood of achieving successful reperfusion was similar across demographic and insurance groups. However, stroke severity at admission varied significantly among these factors. Black patients had significantly higher odds of presenting with severe stroke (NIHSS ≥9: OR = 2.10, p = 0.026; mRS 3-6: OR = 2.35, p = 0.019), suggesting that they arrived at the hospital with more severe strokes compared to other racial groups. Similarly, Medicare patients had the highest odds of severe stroke at admission (NIHSS ≥9: OR = 3.45, p < 0.001; mRS 3-6: OR = 3.89, p < 0.001), indicating that these patients experienced greater stroke burden before intervention (Table 6).

**Table 6:** Logistic Regression Analysis Summary (Adjusted Odds Ratios with 95% Confidence Intervals)

| Outcome | Predictor Variable | Adjusted OR | 95% CI | p-value |
| --- | --- | --- | --- | --- |
| Procedural Reperfusion (TICI 2B-3) | Gender (Female) | 1.02 | 0.78 – 1.34 | 0.854 |
|  | Race (Black) | 0.97 | 0.68 – 1.45 | 0.784 |
|  | Insurance (Medicare) | 1.2 | 0.81 – 1.77 | 0.854 |
|  | Insurance (Medicaid) | 1.03 | 0.21 – 5.00 | 0.976 |
|  | Insurance (Commercial) | 0.79 | 0.18 – 3.39 | 0.751 |
| Procedural Reperfusion (TICI 2C-3) | Gender (Female) | 1.08 | 0.79 – 1.45 | 0.703 |
|  | Race (Black) | 1.12 | 0.75 – 1.55 | 0.784 |
|  | Insurance (Medicare) | 1.35 | 0.87 – 2.15 | 0.703 |
|  | Insurance (Medicaid) | 1.46 | 0.61 – 3.51 | 0.4 |
|  | Insurance (Commercial) | 1.07 | 0.50 – 2.32 | 0.855 |
| NIHSS at Admission | Gender (Female) | 1.25 | 0.95 – 1.64 | 0.097 |
|  | Race (Black) | 2.1 | 1.15 – 3.82 | 0.026 |
|  | Insurance (Medicare) | 3.45 | 2.01 – 5.91 | <0.001 |
|  | Insurance (Medicaid) | 1.49 | 0.64 – 3.49 | 0.383 |
|  | Insurance (Commercial) | 1.57 | 0.72 – 3.39 | 0.255 |
| mRS at Admission | Gender (Female) | 1.18 | 0.90 – 1.55 | 0.112 |
|  | Race (Black) | 2.35 | 1.27 – 4.36 | 0.019 |
|  | Insurance (Medicare) | 3.89 | 2.35 – 6.44 | <0.001 |
|  | Insurance (Medicaid) | 2.45 | 1.03 – 5.84 | 0.044 |
|  | Insurance (Commercial) | 1.38 | 0.67 – 2.86 | 0.382 |
| NIHSS at Discharge | Gender (Female) | 1.16 | 0.81 – 1.66 | 0.411 |
|  | Race (Black) | 1.76 | 0.92 – 3.34 | 0.087 |
|  | Insurance (Medicare) | 1.94 | 0.95 – 3.94 | 0.068 |
|  | Insurance (Medicaid) | 1.48 | 0.62 – 3.54 | 0.383 |
|  | Insurance (Commercial) | 1.57 | 0.72 – 3.39 | 0.255 |
| mRS at Discharge | Gender (Female) | 1.52 | 1.10 – 2.08 | 0.036 |
|  | Race (Black) | 3.12 | 1.35 – 6.75 | 0.021 |
|  | Insurance (Medicare) | 6.73 | 3.71 – 12.23 | <0.001 |
|  | Insurance (Medicaid) | 2.05 | 0.83 – 5.05 | 0.12 |
|  | Insurance (Commercial) | 1.46 | 0.68 – 3.10 | 0.331 |
| mRS at 90 Days | Gender (Female) | 1.52 | 1.10 – 2.08 | 0.036 |
|  | Race (Black) | 3.05 | 1.16 – 8.00 | 0.024 |
|  | Insurance (Medicare) | 6.61 | 3.17 – 13.80 | <0.001 |
|  | Insurance (Medicaid) | 1.79 | 0.62 – 5.20 | 0.283 |
|  | Insurance (Commercial) | 1.6 | 0.61 – 4.19 | 0.334 |
| **Discharge Disposition** | Gender (Female) | 1.56 | 0.95 – 2.55 | 0.077 |
|  | Race (Black) | 1.05 | 0.47 – 2.35 | 0.899 |
|  | Insurance (Medicare) | 5.18 | 1.21 – 22.25 | 0.027 |
|  | Insurance (Medicaid) | 7.31 | 1.53 – 34.97 | 0.013 |
|  | Insurance (Commercial) | 1.62 | 0.33 – 7.99 | 0.556 |
| **tPA Administration** | Gender (Female) | 1.06 | 0.74 – 1.52 | 0.731 |
|  | Race (Black) | 0.88 | 0.46 – 1.70 | 0.709 |
|  | Insurance (Medicare) | 0.65 | 0.34 – 1.24 | 0.19 |
|  | Insurance (Medicaid) | 0.55 | 0.24 – 1.27 | 0.16 |
|  | Insurance (Commercial) | 1.38 | 0.68 – 2.78 | 0.37 |

All models used logistic regression adjusted for gender, race (reference = White), and insurance (reference = Uninsured).

At discharge and at 90 days, disparities in functional outcomes persisted. Black patients had significantly higher odds of poor functional recovery (mRS 3-6 at discharge: OR = 3.12, p = 0.021), while Medicare patients had the strongest association with worse outcomes at both discharge and 90 days (mRS 3-6: OR = 6.73, p < 0.001). Gender differences were also observed, as female patients had significantly worse functional outcomes compared to males (mRS 3-6 at discharge: OR = 1.52, p = 0.036; at 90 days: OR = 1.52, p = 0.036). Additionally, disparities in discharge disposition were noted, with Medicare and Medicaid patients having significantly higher odds of being discharged to non-home settings (Medicare OR = 5.18, p = 0.027), indicating a greater likelihood of requiring skilled nursing or hospice care post-stroke. These findings highlight substantial differences in stroke severity at presentation and long-term recovery, particularly among Black and Medicare patients, despite comparable procedural success rates across all demographic and insurance groups (Table 6).

## Discussion

In this retrospective study of acute ischemic stroke patients undergoing endovascular thrombectomy, we examined whether race, gender, and insurance status influenced treatment success and clinical outcomes. Our findings demonstrate that technical outcomes of EVT are equivalent across different race, gender, and insurance groups, but significant disparities persist in stroke severity at presentation and in post-stroke functional recovery. These results refine our understanding of where and how outcome gaps occur in the stroke care continuum.

We found no significant differences in reperfusion success rates by race, gender, or insurance. All groups achieved a high rate of successful recanalization (∼85–90%), and logistic regression confirmed that patient demographics did not influence the likelihood of achieving reperfusion. This is an important and encouraging finding, indicating that once patients reach the point of receiving endovascular therapy, the quality and efficacy of the procedure are comparable. Our results align with prior studies that reported similar technical and short-term outcomes between racial groups after thrombectomy.^9^ For example, Bouslama et al. found no difference in successful reperfusion or 90-day outcomes between Black and White stroke patients treated with EVT at a high-volume center.^9^ Our study extends this equity in care to insurance status as well, suggesting that within a comprehensive stroke center setting, providers can achieve uniformly high technical success irrespective of a patient’s socioeconomic background or insurance coverage. This likely reflects standardized protocols and technical expertise that have become widespread in the thrombectomy era, as well as a commitment to guideline-driven care that does not vary by patient demographics.^6,12^

### Disparities in stroke severity at presentation

Despite similar in-hospital treatment, patients from different demographic groups did not start on equal footing. Black patients and those with Medicaid or no insurance had significantly higher NIHSS scores on arrival, indicating more severe strokes. There are several potential explanations for this finding. First, the impact of the social determinants of health in these populations is well known.^13,14^ These populations may have decreased access to preventive care resulting in higher burden of stroke risk factors (e.g., hypertension, diabetes) that predispose to more severe strokes or large vessel occlusions.^9^ Our data showed higher rates of hypertension and diabetes in Black patients, consistent with national statistics on risk factor disparities. Second, differences in pre-hospital factors such as recognition of stroke symptoms and delays in seeking care could play a role. Prior studies have noted that certain groups, including Black women, are more likely to arrive outside of optimal treatment windows for tPA,^1^ often due to systemic barriers and lack of awareness.^13,15^ While our study focused on patients who ultimately received EVT (often requiring them to present within a viable time window), it is plausible that lack of access to primary stroke centers led to higher stroke severity by the time of treatment for some patients.^16–18^ Uninsured individuals, for example, might hesitate to seek care early due to cost concerns or lack of primary care, resulting in more extensive brain injury when they do arrive. Our finding that uninsured and Medicaid patients were the youngest on average yet had very high NIHSS suggests that socioeconomic factors, not age or comorbidity alone, contribute to stroke severity in these groups. This is supported by other studies showing uninsured patients tend to have greater neurologic impairment at presentation.^3^ These disparities in initial severity are critical, because initial NIHSS is one of the strongest determinants of outcome, as our analysis and many others have shown.

### Disparities in post-acute functional recovery

The most striking gaps in our study emerged in longer-term functional outcomes. Black and underinsured patients were significantly less likely to achieve functional independence (mRS 0–2) at 90 days compared to White and insured patients, even though their rates of reperfusion were similar. However, once we adjusted for baseline NIHSS (and other factors), race and insurance were no longer independent predictors of outcome, implying that a large part of the outcome disparity is mediated by stroke severity at onset. In plain terms, because Black and underinsured patients had more severe strokes to begin with, they naturally had worse outcomes on average. Yet, our data also suggest that differences in post-stroke care may compound this effect. We observed that disadvantaged patients were less likely to be discharged to rehabilitation facilities and more likely to go to nursing homes or have limited home health services, even when controlling for stroke severity. This finding resonates with recent reports highlighting inequities in access to post-acute stroke rehabilitation based on race and insurance. A study by Man and colleagues found that uninsured stroke patients had far lower odds of being discharged to inpatient rehab compared to those with private insurance.^19^ Reduced access to intensive rehabilitation and support can adversely affect functional recovery, potentially widening the gap in outcomes over time.^20^ Thus, the poorer longer-term outcomes in Black and underinsured patients in our study likely reflect a combination of more severe initial injury and less robust recovery resources.

It is noteworthy that in our cohort, sex differences in outcome were minimal after accounting for age. Women did not have significantly different adjusted outcomes than men, aside from being older at stroke onset. This contrasts somewhat with historical data suggesting women have worse stroke outcomes,^1^ but our finding may be due to improvements in acute care and the fact that all patients in our study received EVT, potentially mitigating some differences. Another consideration is that we did not find a significant sex difference in reperfusion or treatment times (apart from a slight delay in door-to-puncture for women, which could be due to various factors such as vascular access challenges or need for additional consent from family if cognitive issues). The lack of difference in outcomes by sex in our study is reassuring and suggests that when women receive equal advanced therapies, their prognosis can be comparable to men’s, in line with some recent thrombectomy series that also found no sex disparity in outcomes.^21–24^

### Clinical and policy implications

Our findings have important implications for addressing stroke disparities. The fact that the acute intervention itself is delivered equitably and with equal success is heartening; it suggests that stroke centers can provide high-quality care regardless of patient background, and that conscious or unconscious bias at the point of treatment is minimal in this setting. However, the battle is not won at reperfusion alone. The front end (prior to hospital) and back end (post-acute care) of stroke treatment appear to be where disparities arise. To reduce the higher stroke severity observed in Black and underinsured patients, we need community-level and pre-hospital interventions. These could include targeted public education in high-risk communities about stroke symptoms and the importance of early EMS activation, improved access to primary care and blood pressure control in underserved populations, and policies that reduce barriers to seeking emergency care (for instance, expanding insurance coverage or community health worker programs to encourage early hospital arrival). Prior work emphasizes that late hospital arrival is a major reason for treatment exclusion in minorities;^1^ thus, interventions such as community stroke screening events, campaigns in minority media, and partnerships with local organizations could help close the gap in arrival times and initial stroke severity.

On the post-acute side, our study underscores the critical need for equitable access to rehabilitation and follow-up services. Patients without adequate insurance often face difficulties in obtaining intensive inpatient rehab or skilled home therapy, which can limit recovery. Policy measures to address this might include expanding Medicaid programs or subsidy initiatives to cover rehabilitation for uninsured patients, enforcing acute care protocols that arrange rehab for all suitable stroke patients regardless of payor, and developing stroke recovery programs through community clinics for those who cannot access standard rehabilitation. The disparities in discharge disposition we observed support calls for healthcare reforms ensuring that discharge planning is based on clinical need rather than insurance status.^19^ In addition, healthcare systems could invest in stroke navigator programs or case managers who specifically focus on connecting underinsured patients to outpatient therapies, stroke education, and social support to improve recovery trajectories.

Another implication is the potential role of stroke severity as a quality metric: If certain groups consistently present with higher NIHSS, it may reflect broader societal inequities in health. Efforts outside the hospital, such as better hypertension management in Black communities or improved access to preventive care for the uninsured, are essential. Our data suggest that equalizing stroke severity at presentation across groups would substantially equalize outcomes, given the equal effectiveness of EVT we observed. This lends support to multi-faceted public health approaches to stroke prevention and emergency response in disadvantaged populations.^13,25,26^

### Insurance status and functional outcomes

Although Medicare is typically categorized as “insured”, our data show that Medicare patients had significantly worse stroke severity at admission and worse functional outcomes at 90 days. This likely reflects factors tied to age, such as baseline disability, frailty, and a higher burden of comorbidities, rather than access to care alone.^27,28^ These findings challenge the assumption that being “insured” inherently equates to better outcomes. In this context, Medicare status may act as a proxy for clinical vulnerability, and grouping Medicare with private insurance may obscure important disparities in stroke outcomes.

### Comparison with previous studies

The literature on disparities in EVT outcomes is evolving. Earlier studies focusing on racial differences in thrombectomy outcomes have been somewhat inconsistent. Some, as mentioned, showed no significant differences in outcome between racial groups,^9^ while others have noted that after adjusting for variables like stroke severity, race was not an independent predictor of outcome – similar to our findings. For instance, studies have found that Black and Hispanic patients had worse unadjusted functional outcomes after thrombectomy compared to Whites, but these differences were largely accounted for by differences in baseline factors, with no race effect in multivariable models – aligning with our conclusions.^29–31^ Gender differences in EVT outcomes have also been explored; many studies report that while women may have worse raw outcomes, sex is not an independent predictor after accounting for age and comorbidities,^22,32,33^ which our study corroborates. With respect to insurance, literature is abundant on general stroke outcomes such as uninsured patients having higher mortality and longer length of stay,^2,3,34^ but specific data on EVT populations are scarcer. Our study contributes by showing that within an EVT-treated cohort, insurance status correlates with outcome primarily through pathways of initial severity and post-acute care rather than intra-procedural differences.

### Strengths and limitations

A strength of our study is the comprehensive inclusion of multiple disparity axes (race, gender, insurance) in one cohort, allowing us to examine their interrelated effects. Our sample, drawn from a high-volume stroke center serving a diverse urban population, included substantial numbers of patients from traditionally underrepresented groups (nearly 40% Black, and ∼25% underinsured), enhancing the relevance of our findings to real-world practice. We also had systematic follow-up for functional outcomes and granular data on process measures, enabling a deeper analysis of where differences arose.

However, several limitations warrant consideration. First, as a single-center retrospective study, our findings may not generalize to all settings, particularly rural or low-resource hospitals. The equitable procedural outcomes we observed could reflect the culture and protocols of our center; other systems might have different results. Second, the categorization of race and ethnicity was broad; we did not analyze differences between, for example, Hispanic subgroups or other minorities due to smaller numbers. Third, insurance status was used as a proxy for socioeconomic status, but it may not capture all dimensions of disadvantage—factors like income, education, or neighborhood characteristics (social determinants of health) were not directly measured. These unmeasured factors could contribute to both stroke severity and recovery disparities. Fourth, while we identified associations with discharge disposition, our study did not directly measure the quality or intensity of rehabilitation each patient received, which would be valuable in understanding the recovery gap. Finally, there could be selection bias in that all patients in this study survived long enough and were deemed candidates for EVT; those with extremely severe strokes who died early or those never transferred for thrombectomy might have differed by demographic factors, which we could not assess. Despite these limitations, our study provides meaningful insights by leveraging newly updated data to clarify that disparities in stroke outcomes after EVT are largely rooted in pre- and post-hospital factors rather than differences in the emergency treatment itself.

### Future directions

Addressing the identified disparities will require both medical and societal efforts. Future research should explore interventions aimed at reducing pre-hospital delays in minority communities – for example, evaluating the impact of community health worker outreach or mobile stroke units in underserved areas on reducing time to treatment and stroke severity. Additionally, studies could assess post-acute intervention programs (such as providing subsidized rehab or tele-rehabilitation for underinsured patients) to see if the functional outcome gap can be narrowed. From a research standpoint, incorporating more detailed socioeconomic and geographic data into stroke registries will help disentangle the various contributors to outcome disparities. On a broader scale, advocacy for health policy changes, such as expanding Medicaid in states that have not done so and improving insurance coverage for comprehensive stroke care, is supported by evidence like ours that such factors do affect outcomes. Eliminating disparities in stroke outcomes is a multifactorial challenge, but our findings highlight specific leverage points: ensuring timely, high-quality acute treatment for all, which we show is achievable, and bridging the gaps in stroke severity through prevention and in recovery through equitable rehabilitation access.

## Conclusion

This study demonstrates that while procedural success rates for EVT were equitable across gender, racial, and insurance groups, significant disparities persisted in stroke severity at admission and long-term functional recovery. Black and Medicare patients presented with more severe strokes, which contributed to their worse functional outcomes at discharge and 90 days. Despite similar procedural reperfusion success, Medicare and Medicaid patients had significantly higher odds of poor functional recovery and were more likely to be discharged to non-home settings. Female patients also experienced higher post-stroke disability than males, despite similar EVT success rates.

These findings suggest that disparities in stroke outcomes are largely driven by pre-hospital factors, including delays in seeking care and differences in baseline stroke burden, as well as post-hospital barriers to rehabilitation access. Addressing these inequities will require targeted interventions beyond the hospital setting, including expanded community-based stroke awareness programs, improved EMS response times, and increased access to preventive stroke care. Additionally, health policy reforms aimed at improving post-stroke rehabilitation access for Medicare and Medicaid patients will be essential to reducing long-term disparities in functional recovery.

From a hospital perspective, the consistent EVT success rates across all groups indicate that stroke care is being delivered equitably. However, hospitals must also play a role in improving stroke outcomes by strengthening discharge planning, facilitating rehabilitation referrals, and advocating for systemic healthcare improvements. Moving forward, a multidisciplinary approach integrating hospital-based excellence with community-level interventions and healthcare policy changes will be essential to ensuring more equitable stroke treatment and recovery for all patients.

